# Predicting thermal injury patient outcomes in a Tertiary-Care Burn Centre, Pakistan

**DOI:** 10.1101/2021.06.14.21258932

**Authors:** Mohammad Suleman Bajwa, Muhammad Sohail, Hamza Ali, Umer Nazir, Muhammad Mustehsan Bashir

**Affiliations:** Mayo Hospital, Lahore, Pakistan; Mayo Burn Centre/Plastic & Reconstructive Surgery Department, Mayo Hospital, Lahore

**Keywords:** burn, survival, length of hospital stay, predictive model

## Abstract

**Objectives:** To explore candidate parameters for their ability to predict *survival* and *length of hospital stay (LOS)* in thermal burns patients, to prepare multivariate predictive models for these two outcomes, and to compare performance of native models to other models.

**Methods:** A retrospective cohort study was undertaken based on record review. Data was extracted from files of patients admitted to a tertiary-care burn centre in Lahore, Pakistan from January 1, 2020 to October 31, 2020. Univariate preselection was used to prepare multivariate logistic regression models for each outcome of interest (survival and LOS). Multivariate models were tested and compared to other models.

**Results:** Increasing TBSA of burn was positively associated with reduced survival and prolonged length of stay. Advancing age and full-thickness burns independently predicted decreased survival. Burn etiology showed prognostic value: petrol-flame burns predicted decreased survival and prolonged LOS; scald was associated with improved survival-odds and shorter LOS. The Survival-model consisted of (1) Baux score, (2) TBSA>40% and (3) serum albumin <3.5g/dl (AUC=0.968, Nagelkerke R^2^=0.797). The LOS-model consisted of (1) TBSA^**2**^ and (2) serum albumin concentration (AUC= 0.832, Nagelkerke R^2^ =0.408). In tests of discrimination and calibration, native models prepared for survival and LOS outcomes outperformed other models applicable to our dataset.

**Conclusion:** Data from a South Asian burn center has been used to explore factors influencing prognosis for their utility in predictive models for survival and the duration of hospital stay. The significant prognostic roles of TBSA, age, inhalational injury, burn-depth, etiology of burn, anatomic site of burn, hypoalbuminemia and other biochemical parameters were observed. These tools hold significance in guiding healthcare policy and in communications with patients and their families.

## Introduction

Models and scoring indices predictive of burn patient outcomes are quality improvement tools. Efficient predictions of survival and length of hospital stay (LOS) can support effective triage, allow for better communication of expectations with patients and their families, guide healthcare policy and help individualize patient management, including decisions of withholding or withdrawing therapy^[1]^. A variety of predictive models have been proposed^[2,3]^. The role of popular scoring indices, including BOBI^[4]^, ABSI^[5]^, Baux score^[6]^, Revised Baux score^[7]^ and the estimate of LOS being 1day/%TBSA (total body surface area, burnt)^[8]^, in predicting outcomes has been questioned^[2]^. Consensus in prediction is difficult, and is attributed to differences in patient populations and management protocols^[9]^. It has been recommended that future models explore less-studied variables for their predictiveness to better account for variations^[2]^. As a result, the role of biochemical parameters in prediction has recently gained recognition^[10-12]^.

With the South-Asian population being under-represented in predictive models^[13]^, published predictive tools require validation in this population. The unique characteristics of South-Asian burn patients and their care; the high incidence of burns^[14]^, undeveloped burn prevention systems and other poverty-related circumstances as malnutrition, poor living conditions, use of unsafe stoves^[15]^, and inadequate access to healthcare facilities suggest the need for the development of local models which may hold greater validity for this population. Models developed and updated in individual burn centers may be of greater clinical utility compared to models developed in different settings^[9]^.

The authors conducted this study to explore clinical and biochemical parameters predictive of our two outcomes of interest (survival and LOS) in thermal burn patients admitted to a South Asian burn center, and to prepare a predictive model for each outcome. The predictive performance of these native models, published large-scale predictive models and models derived from scoring indices in our population were studied and compared.

## Methods

IRB approval was secured to conduct a retrospective cohort study based on record review. Thermal burn patients admitted to the Burn Centre, Mayo Hospital, Lahore from January 1, 2020 to October 31, 2020 were included. Non-thermal burns, re-admissions, incomplete records, those who were discharged on request, left against medical advice, or who presented later than 24 hours of being burnt were excluded. Our burn centre’s admission criteria for thermal burn patients includes all patients with partial-thickness burns of 15% TBSA or greater, patients with full-thickness or deep wounds of any surface area, patients with significant co-morbidities, and those with burns involving face, hands, feet, genitalia, perineum or major joints in need of specialized rehabilitative intervention. Patients with burns of greater than 30% TBSA, inhalation injury, known comorbidities (e.g., diabetes mellitus, hypertension, chronic kidney disease) and/or concerning signs of systemic inflammation are managed in the Intensive Care Unit (ICU). We defined length of hospital stay (LOS) as the period between hospital admission and hospital discharge. Inhalational injury is defined by presence of facial burns with singed nasal hair, carbonaceous sputum, stridor or laboured breathing, or history of a closed-space fire. ‘Dressing-change under general anesthesia’ is considered an operative intervention in this study. Patients with burns predominantly of superficial-partial thickness were managed conservatively with interactive dressings.

Data was physically extracted from patient files. Parameters recorded include patient age and gender, clinical parameters (TBSA, wound depth, anatomical site of burn, etiology of burn), comorbidities, biochemical parameters within 24 hours of presentation (serum electrolytes, albumin, ALT, urea, creatinine, TLC, Platelet count, PT, aPTT, INR), ICU care, operative management, survival and LOS of patients. ABSI, Baux score, revised-Baux score and other variables of published models were computed. ABSI has been stratified into a 6-point scale^[5]^, which we labeled ‘ABSI risk-stratified’. Burn depth in patients is described according to the predominant depth of burns, according to which the management strategy was tailored. Cases were coded for anonymity.

SPSS version 23 was used for statistical analysis, described in *figure 1*. Single imputation was used to ‘fill in’ the missing data which was limited to the variables of serum ALT (14% missing data) and coagulation profiles (6.3% missing data). Regression was used to impute, while employing constraints of minimum and maximum values, as it serves to preserve the relationship between predictors and variables^[16]^.

**Figure 1:**
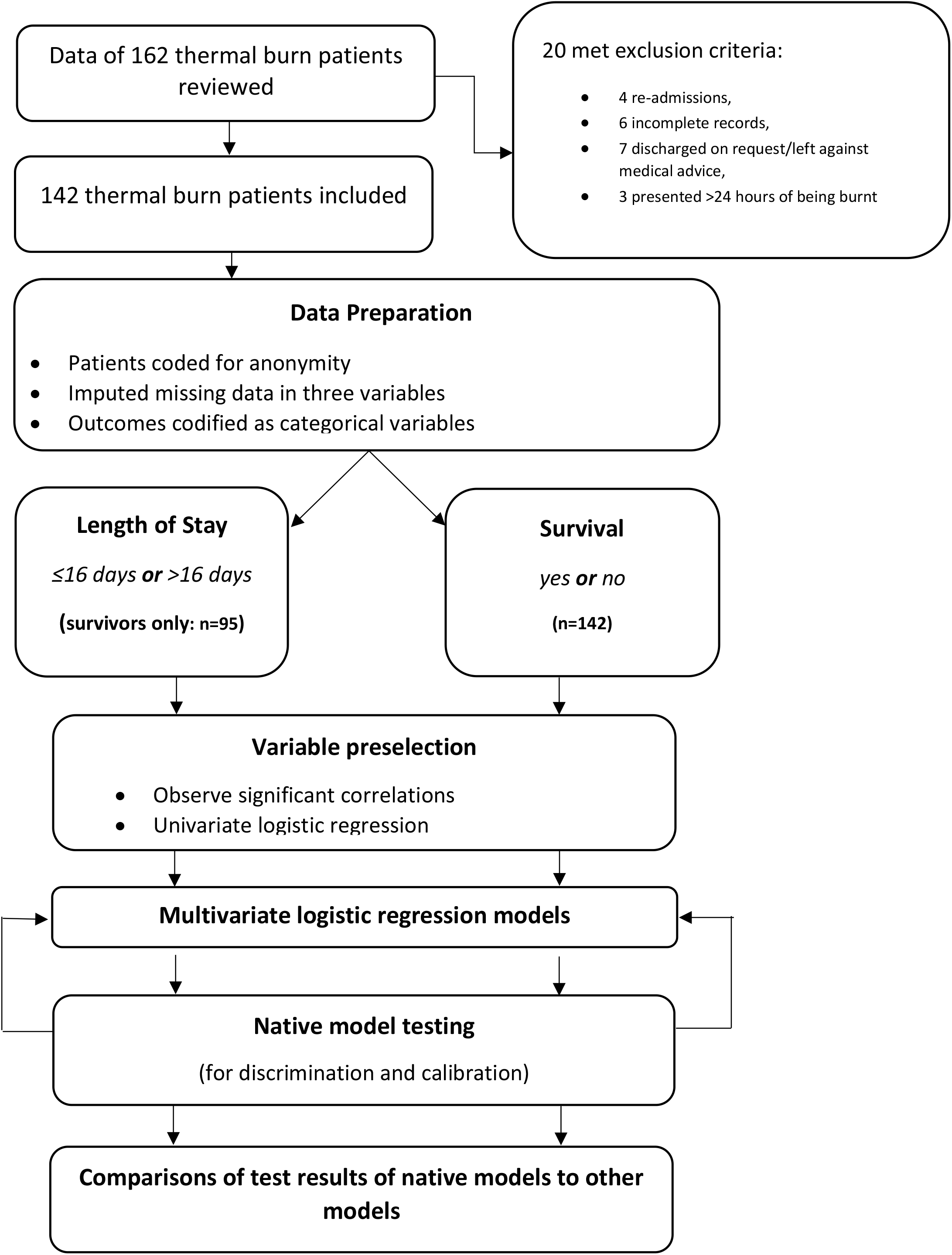
Flowchart of methodology.

In order to code LOS into a nominal variable, its distribution in survivors was tested for normality, to decide on a measure of central tendency. LOS was not normally distributed so the median (LOS=16 days) was used to divide patients into categories: short LOS/early recovery (≤16 days), and prolonged LOS/late recovery (>16 days).

Similar steps were used in preparing the Survival-predictive model and the LOS-predictive, although the LOS-predictive model was developed using data of surviving patients only. Significant correlations between covariates and each outcome were observed, while screening for multicollinearity.

Univariate logistic regression was then employed for variable preselection. Odds ratios and p-values were reported. As certain parameters (operative management, LOS, ICU LOS) would not be known on day 1 of presentation, they were excluded from multivariate modeling. Only biologically plausible factors of p<0.2 were selected for multivariate logistic regression. Models were prepared using forward selection and backwards elimination based on the Akaike Information Criterion (AIC), arriving at the most predictive and significant models after multiple trials. To avoid ‘overfitting’, we ensured at least 10 outcome events were present for each variable in the final model^[17]^. Performance of the models was assessed by Receiver Operating Characteristic (ROC) curve analysis and Hosmer-Lemeshow (HL) ‘goodness-of-fit’ test. The odds ratios, regression coefficients, p-values and pseudo-R^[2]^ values of the models were reported.

## Results

We included 142 patients in the final analysis. *Table 1* describes patient demographics. Ninety-five (66.9%) patients survived. Patient with predominantly superficial-partial burns all survived (p=0.008), while only 8(40%) of those with predominantly full-thickness burns survived (p=0.008). The median length of hospital stay (LOS) was 16 days (IQR=22). Early recovery (≤16 days) had significant positive correlation to superficial-partial burn patients (p=0.023), and late recovery (>16 days) with predominant deep-partial burns(p=0.003). The mean TBSA was 32.68% ± 25.3, with positive correlation of TBSA to full-thickness burns(p=0.023) and negative correlation to superficial partial burns(p=0.001). Eighty-seven (61.3%) patients who underwent surgery were significantly positively correlated(p<0.05) to deep-partial thickness burns and higher levels of serum albumin on the first day of admission, and had significant negative correlations (p<0.05) to advancing age, increasing TBSA, superficial-partial burns, full-thickness burns, buttock burns, perineal burns and hypokalemia. Twenty-six (18.3%) patients were admitted to the ICU and showed significant positive correlation to increasing TBSA, lower limb burns, back burns, hypoalbuminemia and to being victims of cylinder-blast burns (p<0.05).

**Table 1.**
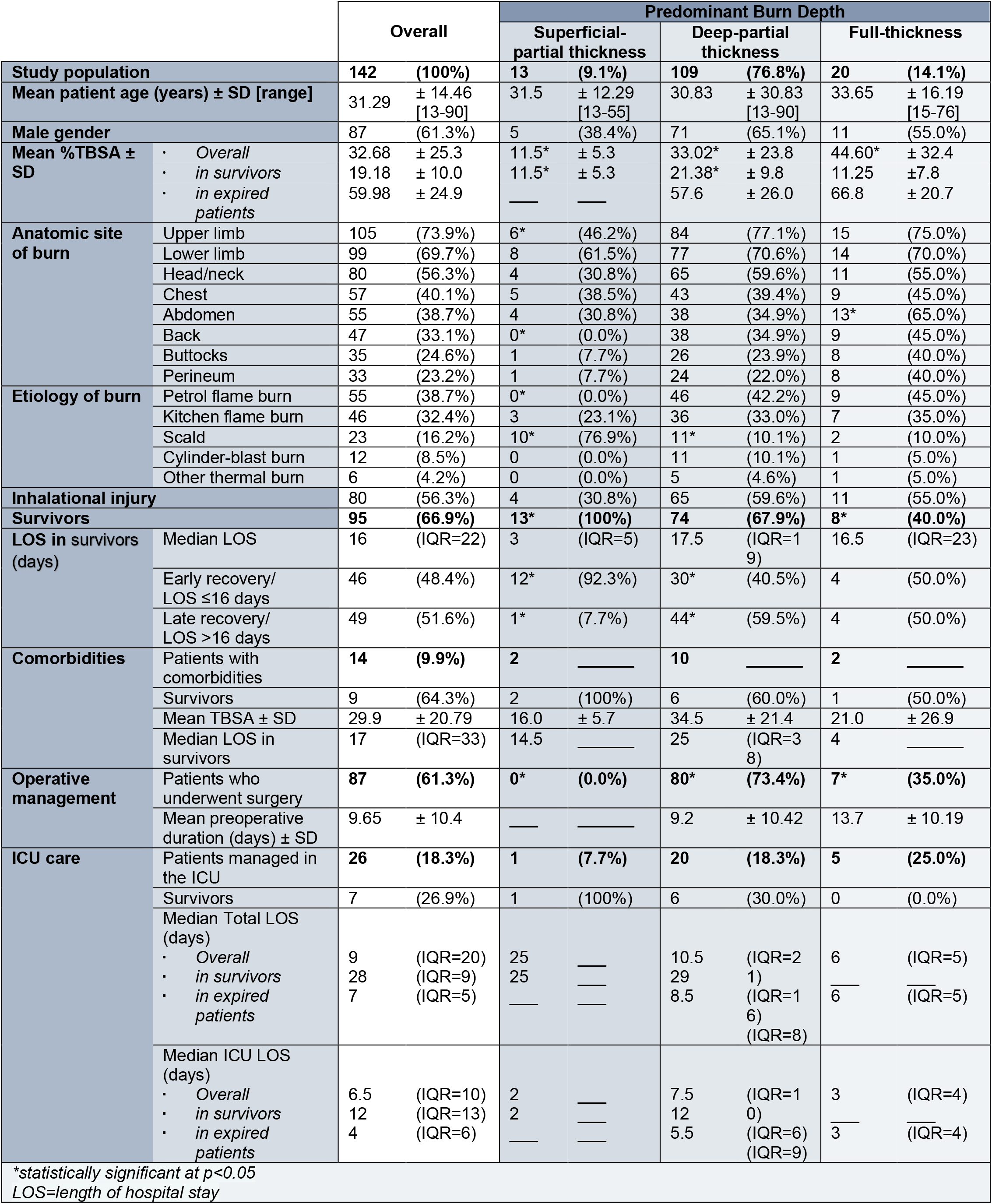
Patient demographics.

### Univariate analysis results

Variables underwent univariate preselection for each model. Odds ratios for variables of survival and LOS, having significance of p<0.2, are reported in *Tables 2 and 3*, respectively. For every 10% increase in TBSA, the odds of survival reduced by 3.7 times (CI=2.38-5.88, p=0.001) and the odds of long LOS in burn survivors increased by 3.4 times (CI=1.93-5.97, p=0.00).

**Table 2:** Univariate regression results for survival.

| Variable |  | Odds ratio | Confidence Interval | p value |
| --- | --- | --- | --- | --- |
| LOS |  | 1.137 | 1.075-1.202 | 0.00 |
| Age | (-) | 0.95 | 0.929-0.980 | 0.00 |
| Age(in decades) | (-) | 0.61 | 0.470-0.786 | 0.00 |
| TBSA | (-) | 0.87 | 0.824-0.911 | 0.00 |
| TBSA<sup>2</sup> | (-) | 0.99 | 0.997-0.999 | 0.00 |
| TBSA (in deciles) | (-) | 0.27 | 0.170-0.420 | 0.00 |
| TBSA >40 | (-) | 0.01 | 0.003-0.036 | 0.00 |
| Baux score | (-) | 0.90 | 0.863-0.932 | 0.00 |
| R-Baux score | (-) | 0.94 | 0.922-0.961 | 0.00 |
| ABSI | (-) | 0.38 | 0.268-0.531 | 0.00 |
| ABSI risk-stratified | (-) | 0.17 | 0.095-0.303 | 0.00 |
| BOBI | (-) | 0.49 | 0.387-0.625 | 0.00 |
| No. of operations |  | 7.37 | 3.419-15.908 | 0.00 |
| Operative management |  | 35.81 | 8.198-156.437 | 0.00 |
| Petrol flame | (-) | 0.40 | 0.195-0.825 | 0.01 |
| Scald |  | 3.67 | 1.027-13.096 | 0.05 |
| ICU care | (-) | 0.12 | 0.045-0.308 | 0.00 |
| ICU LOS | (-) | 0.88 | 0.798-0.976 | 0.15 |
| Full-thickness burn | (-) | 0.27 | 0.101-0.712 | 0.01 |
| Chest burn | (-) | 0.12 | 0.055-0.270 | 0.00 |
| Abdominal burn | (-) | 0.20 | 0.096-0.428 | 0.00 |
| Trunk burn | (-) | 0.05 | 0.018-0.137 | 0.00 |
| Upper limb burn | (-) | 0.23 | 0.084-0.650 | 0.01 |
| Lower limb burn | (-) | 0.35 | 0.148-0.837 | 0.02 |
| All limbs burnt | (-) | 0.07 | 0.030-0.176 | 0.00 |
| Back burn | (-) | 0.11 | 0.047-0.234 | 0.00 |
| Perineal burn | (-) | 0.08 | 0.032-0.204 | 0.00 |
| Buttocks burn | (-) | 0.13 | 0.054-0.294 | 0.00 |
| Serum potassium >5mEq/L | (-) | 0.44 | 0.167-1.134 | 0.09 |
| Serum potassium <3.5 or >5mEq/L | (-) | 0.59 | 0.264-1.314 | 0.20 |
| Serum Urea>20mg/dl | (-) | 0.33 | 0.106-1.018 | 0.05 |
| Creatinine (>1.3 mg/dl in males, >1.1 mg/dl in females) | (-) | 0.23 | 0.094-0.555 | 0.00 |
| Albumin <3.5g/dl | (-) | 0.26 | 0.107-0.647 | 0.00 |
| Serum ALT >40 IU/L | (-) | 0.61 | 0.297-1.232 | 0.17 |
| *(-) represents a negative predictor* |  |  |  |  |

**Table 3:** Univariate regression results for LOS.

| Variable | Odds ratio | Confidence Interval | p value |
| --- | --- | --- | --- |
| TBSA | 1.16 | 1.086-1.237 | 0.00 |
| TBSA<sup>2</sup> | 1.004 | 1.002-1.006 | 0.00 |
| TBSA (in deciles) | 3.4 | 1.931-5.968 | 0.00 |
| Baux score | 1.04 | 1.013-1.076 | 0.01 |
| R-Baux score | 1.03 | 1.003-1.053 | 0.03 |
| ABSI | 1.62 | 1.214-2.173 | 0.00 |
| ABSI risk-stratified | 2.91 | 1.617-5.227 | 0.00 |
| BOBI | 1.19 | 0.933-1.505 | 0.17 |
| No. of operations | 3.87 | 1.767-8.459 | 0.00 |
| Days till first operation | 1.24 | 1.072-1.444 | 0.00 |
| Operative Management | 21 | 2.630-167.671 | 0.00 |
| Petrol flame | 3.28 | 1.265-8.485 | 0.02 |
| Scald (-) | 0.18 | 0.056-0.606 | 0.01 |
| Male gender | 1.72 | 0.753-3.984 | 0.20 |
| Sup.-partial thickness burn (-) | 0.06 | 0.007-0.476 | 0.01 |
| Deep-partial thickness burn | 4.69 | 1.553-14.19 | 0.01 |
| Upper limb burn | 1.95 | 0.821-4.624 | 0.13 |
| Lower limb burn | 2.54 | 1.076-5.989 | 0.03 |
| All limbs burnt | 10.13 | 1.228-83.466 | 0.03 |
| Back burn | 2.37 | 0.755-7.463 | 0.14 |
| Perineal burn | 3.07 | 0.587-16.057 | 0.18 |
| Albumin<3.5g/dl | 2.73 | 1.168-6.367 | 0.02 |
| TLC>11,000 WBC/mm<sup>3</sup> | 1.98 | 0.822-4.779 | 0.13 |
| (-) represents a negative predictor |  |  |  |

Survival in burn patients (see *Table 2*) was positively predicted by prolonged LOS and surgical management and negatively predicted by mortality indices, requirement of ICU care and presence of full-thickness burn. Petrol flames decreased survival-odds by 2.5 times, in contrast to scald burns which increased odds of survival 3.7 times in admitted patients. Burns involving perineum or large anatomical regions as the anterior trunk, or four limbs were the strongest negative predictors of survival from amongst anatomical sites. Raised levels of serum potassium, urea and creatinine, and hypoalbuminemia were significant negative predictors of survival.

Prolonged LOS in burn survivors (see *Table 3*) was positively predicted by mortality indices as Baux and Revised-Baux (R-Baux) score, ABSI and BOBI. Surgical management, particularly that involving multiple operations, increased odds of long hospital stay. Petrol flames increased odds of late recovery by 3.3 times, while scald burns increased odds of early recovery 5.5 times. Perineal, back and extremity burns were strong predictors of long hospital stay. Hypoalbuminemia and leukocytosis were positive predictors of long LOS. We observed male patients to have increased odds ratio for long LOS.

### Multivariate analysis results

The Survival-predictive model *(Table 4)* correctly classifies 92.3% of cases, compared to a control of 66.9%. Its sensitivity is 95.8% and its specificity is 85.1%. Nagelkerke R2 is 0.797 and Hosmer-Lemeshow test has a chi-square value of 1.82 and p=0.99. The AUC is 0.97 (95% CI is 0.943-0.994). Baux score is the most significant predictor in this model. Using Baux score as a covariate showed greater predictive efficiency, discrimination and calibration compared to expanded mortality indices as ABSI, BOBI, and the R-Baux score. This suggests that the common factors, i.e., age and TBSA are good predictors of survival.

**Table 4:** Survival-predictive model.

| Covariate | Coefficient | Odds Ratio | Confidence Interval | p value |
| --- | --- | --- | --- | --- |
| Intercept | 11.642 | - | - | - |
| Baux score | -0.078 | 0.925 | 0.886-0.966 | 0.000 |
| TBSA>40% | -2.802 | 0.061 | 0.011-0.342 | 0.001 |
| Serum Albumin < 3.5g/dl | -1.927 | 0.146 | 0.022-0.984 | 0.048 |

The LOS-predictive model *(Table 5)* correctly classifies 73.7% of cases compared to a control of 51.6%. It has a sensitivity of 69.4% and specificity of 78.3%. Nagelkerke R^2^ is 0.408 and Hosmer-Lemeshow test has a chi-square value of 5.87 and p=0.56. The area under receiver operating characteristic curve (AUC) is 0.832 (CI=0.749-0.914). TBSA^2^ is the most significant predictor in this model.

**Table 5:**
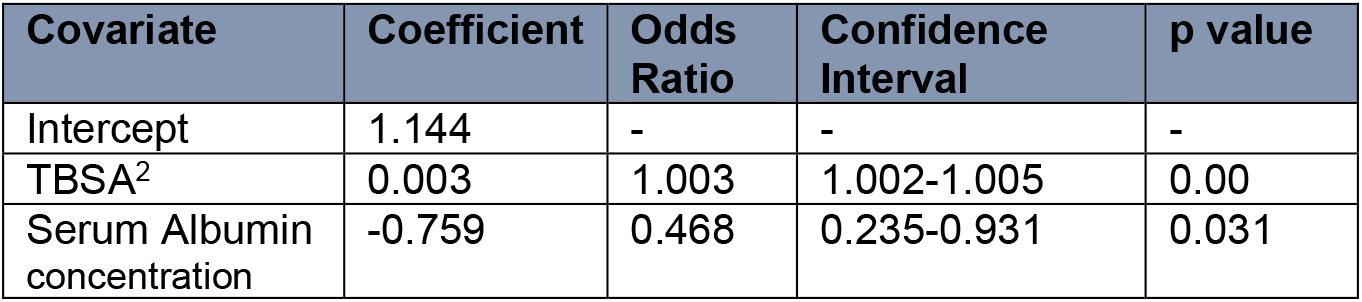
Length of hospital stay – predictive model.

| Covariate | Coefficient | Odds Ratio | Confidence Interval | p value |
| --- | --- | --- | --- | --- |
| Intercept | 1.144 | - | - | - |
| TBSA<sup>2</sup> | 0.003 | 1.003 | 1.002-1.005 | 0.00 |
| Serum Albumin concentration | -0.759 | 0.468 | 0.235-0.931 | 0.031 |

We evaluated the fit of universal models to or native dataset. The results are summarized in these *Tables 6 and 7* below. The variables used by Ho et al^[9]^ in their Survival-model, i.e., TBSA^2^, age^2^ and inhalational injury, were used to make a model which was calibrated to our study population. This model performed better in tests compared to their original Survival-model. Similarly, the LOS-model developed by Ho et al^[9]^, which comprises TBSA, TBSA^2^, inhalational injury and gender, performed better when recalibrated to our population. While ABSI and Baux score were observed to be adequate predictors of LOS and survival, BOBI was a poor predictor of each outcome.

**Table 6:** Comparison of Survival-predictive models.

|  | No. of variables | %correctly predicted (sensitivity/specificity) | Hosmer-Lemeshow test Chi-square [significance] | AUC | Nagelkerke R<sup>2</sup> |
| --- | --- | --- | --- | --- | --- |
| Null model | 0 | 66.9 (100.0/0.0) | - | - | - |
| *Native model* | 4 | 92.3 (95.8/85.1) | 1.818 [p=0.986]* | 0.968 | 0.797 |
| Ho et al Survival-recalibrated<sup>[9]</sup> | 3 | 91.5 (96.8/80.9) | 6.564 [p=0.584]* | 0.959 | 0.784 |
| Baux score<sup>[6]</sup> ° | 2 | 87.3 (92.6/76.6) | 7.092 [p=0.527]* | 0.949 | 0.726 |
| TBSA of burn | 1 | 90.8 (95.8/80.9) | 12.682 [p=0.123]* | 0.939 | 0.732 |
| ABSI<sup>[5]</sup> ° | 5 | 88.0 (96.8/70.2) | 6.578 [p=0.474]* | 0.928 | 0.691 |
| ABSI- risk-stratified<sup>[5]</sup> ° | 5 | 86.6 (87.4/85.1) | 3.598 [p=0.463]* | 0.914 | 0.666 |
| Ho et al Survival-model <sup>[9]</sup> ° | 3 | 83.8 (93.7/63.8) | 8.187 [p=0.415]* | 0.926 | 0.594 |
| R-Baux score<sup>[7]</sup> ° | 3 | 85.2 (94.7/66.0) | 6.796 [p=0.559]* | 0.916 | 0.548 |
| BOBI<sup>[4]</sup> | 3 | 84.5 (94.7/63.8) | 16.825 [p=0.010] | 0.834 | 0.44 |
| * = significantly-good calibration °=models derived from scoring indices |  |  |  |  |  |

**Table 7:** Comparison of LOS-predictive models.

|  | No. of variables | %correctly predicted (sensitivity/specificity) | Hosmer-Lemeshow test Chi-square [significance] | AUC | Nagelkerke R<sup>2</sup> |
| --- | --- | --- | --- | --- | --- |
| Null model | 0 | 51.6 (100.0/0.0) | - | - | - |
| *Native model* | 2 | 73.7 (69.4/78.3) | 5.87 [p=0.556]* | 0.832 | 0.408 |
| Ho et al LOS-recalibrated<sup>[9]</sup> | 3 | 74.7 (73.5/76.1) | 7.93 [p=0.339]* | 0.824 | 0.395 |
| LOS=1 day per%TBSA<sup>[8]</sup> | 1 | 69.5 (65.3/73.9) | 11.37 [p=0.123]* | 0.812 | 0.371 |
| Ho et al LOS-model<sup>[9]</sup> | 4 | 69.5 (65.3/73.9) | 13.72 [p=0.056]* | 0.809 | 0.356 |
| ABSI risk-stratified<sup>[5]°</sup> | 5 | 66.3 (63.3/69.6) | 0.72 [p=0.697]* | 0.709 | 0.206 |
| ABSI<sup>[5]°</sup> | 5 | 66.3 (63.3/69.6) | 5.32 [p=0.256]* | 0.693 | 0.164 |
| Baux score<sup>[6]°</sup> | 2 | 60.0 (55.1/65.2) | 3.99 [p=0.781]* | 0.664 | 0.12 |
| R-Baux<sup>[7]°</sup> | 3 | 62.1 (65.3/58.7) | 12.07 [p=0.098]* | 0.639 | 0.086 |
| BOBI score<sup>[4]</sup> | 3 | 52.8 (59.2/45.7) | 18.78 [p=0.01] | 0.592 | 0.027 |
| * = significantly-good calibration °=models derived from scoring indices |  |  |  |  |  |

## Discussion

Data from a South Asian burn center was used to explore commonly-recorded clinical and biochemical parameters for their utility as predictors of survival and length of hospital stay (LOS). In agreement with previous studies^[2,3]^, we observed thermal burns of increasing TBSA to predict decreased chances of survival and longer hospital stay, and advancing age to portend fewer chances of survival. Hypoalbuminemia and perineal burns consistently related to decreased survival and prolonged LOS. Female gender showed weak correlation (p=0.2) to LOS but not to survival. Patients with full-thickness burns were significantly associated with increasing TBSA and decreased survival. Early recovery (≤16 days) was significant associated with superficial-partial burn patients, and late recovery (>16 days) with deep-partial burn patients. Burn etiology showed predictive significance in our population. Petrol-flame burns predicted prolonged LOS and decreased survival; scald was associated with shorter LOS and improved survival-odds. Native models prepared for survival and LOS outcomes performed well compared to existing models and models derived from scoring indices applicable to our dataset.

In an effort to improve model performance, different variables have been explored for their prognostic significance. The simplest models use TBSA alone^[8,18]^. The Baux score^[6]^ derives from TBSA and patient age. These factors are joined with inhalational injury in the Revised-Baux score^[7]^, BOBI^[4]^, and the Survival-model of Ho et al^[9]^. The ABSI^[5]^ further includes sex and presence of full-thickness burn. The LOS-model of Ho et al^[9]^ includes TBSA, sex and inhalational injury. Recent studies have explored the prognostic roles of early biochemical findings including hypoalbuminemia^[10,11]^ and abnormal blood total leukocyte count (TLC)^[12]^. The modifiable nature of biochemical parameters enables them to provide greater prognostic insight than many non-modifiable clinical parameters. Studies suggest hypoalbuminemia to be is a strong independent predictor of survival[10] and of LOS^[11]^, in agreement with our observations. A study in Taiwan^[11]^ found hypoalbuminemia positively correlated to early multiorgan dysfunction syndrome, decreased extubation rates and increased ICU LOS. While the prognostic role of early leukocytosis has been suggested^[12]^, we found hypoalbuminemia to hold greater predictive value.

The native Survival-model and LOS-model, both outperformed applicable models in tests of discrimination and calibration. The Survival-model accounted for 96.8% of variation in survival and the LOS-model accounted for 83.2% of variation. This lends to the opinion^[9]^ that burn centers should prepare and update independent models by extracting data from their own population if the objective of a clinically-relevant model is to be achieved. However, incorporation of variables of other models into a local model is an option; recalibration of the models of Ho et al^[9]^ to the study population greatly improved model performance.

Survival showed strong correlation to prolonged LOS and surgical management. Patients who underwent surgical management were about 30 times more likely to survive, which is in agreement with findings of previous studies^[18-20]^. Univariate analysis revealed hypoalbuminemia, hyperkalemia, uremia, high creatinine and raised ALT on admission to be independent negative predictors of survival. While inhalational injury did not meet our criteria for a predictive model of survival, Smith et al^[21]^ found inhalational injury to not be a strong predictor as well. We observed Baux score to be the best predictor of survival from amongst mortality scoring indices.

Length of hospital stay is a difficult outcome to predict; a review^[2]^ found studied models could account for only 15-75% of population variation in LOS. Our univariate regression analysis revealed established mortality indices to be significant predictors of LOS, in agreement with previous studies^[4-6]^. The ‘LOS=1 day/%TBSA’^[8]^ rule was able to account for 81.2% of variation in our population’s LOS, surpassed only by our LOS-model and that of Ho et al^[9]^. We found addition of serum albumin concentration to TBSA, increased predictive efficiency and ability to account for variation in LOS to 83.2%. We observed the anatomic site of burn to be predictive of LOS; burns involving the perineal region significantly correlated to longer LOS and increasing TBSA, similar to the observation of Frugoni et al^[12]^. Burns involving the lower limbs were observed to significantly increase odds of delayed recovery, similar to the observation of Sanderson et al^[19]^. Peripheral artery disease^[22]^, relative vascular insufficiency of large adipose tissue deposits^[23]^, complications of premature weight bearing and of delayed ambulation^[24]^, and poor compliance of patients and family caregivers with the demands of physiotherapy are all limiting factors to healing of lower limb burn wounds. However, we observed a weak significant correlation (p=0.09) of lower limb burns to diagnosed comorbidities in our population; further study is needed to clarify its predictive value. The association of female gender with prolonged LOS has been observed previously^[9,20]^, though we found its impact difficult to assess due to multi-collinearity.

Thermal burn etiology has prognostic significance; we observed scald burn patients had increased odds of survival and early recovery, in contrast to petrol flame burns. However, scalds were significantly (p<0.01) positively correlated to female gender, superficial-partial burns and decreasing burn surface area, while petrol flames showed strongly significant (p<0.01) positive correlation to male gender, deep-partial burns and increasing TBSA. Wong et al^[25]^ found burn etiology predictive of mortality, not LOS, while Bhatia et al^[26]^ found no benefit in adding burn etiology in their model.

As our unit is a public sector tertiary care setting which caters services to densely populated areas of Lahore and its surroundings, patients of low socioeconomic status, who often live and work in hazardous surroundings, are likely to have been over-represented in this study. This population of patients and their family caregivers often cannot bear the indirect costs incurred from prolonged hospitalization, and they choose to be discharged on request or leave against medical advice; economic considerations have a role in predicting LOS. High patient-load attributable to a lack of preventive services, and limitations of beds/justice are also determinants of LOS. Similar problems are faced in other South Asian units^[27, 28]^.

The authors recommend that an improved injury surveillance system, (as the SABR-South Asian Burn Registry^[14]^) be implemented, targeting identified predictive factors and others deemed plausible, in order to better prepare adequate burn prevention strategies such as public education, particularly in rural areas, training of laborers, infrastructural support and implementation of safety standards in kitchen stoves. Predictive models can then be prepared and updated alongside this index system, with separate models for groups suffering burns of different etiologies. Factors less uniformly recorded, including pre-existing health-conditions of patients, patient education, cooperation of family caregivers with treatment and hospital factors are likely to have prognostic significance and warrant investigation in future studies.

### Limitations

This study has some limitations. It observes a selected sample of cases presenting to a single center. Certain variables of interest, as percentage of full-thickness burn, Abbreviated Injury Score, body mass index, history of tobacco use and preadmission steroid use were not consistently available on record-review. The pediatric burns population is not represented; no patients under 13 years’ age were included in this study.

## Conclusion

Data from a South Asian burn center has been used to explore factors influencing prognosis that may be used in locally-relevant predictive models for survival and the duration of hospital stay. The significant prognostic roles of well-studied predictors as TBSA, age, and inhalational injury, as well as burn-depth, biochemical parameters, etiology of burn and anatomic site of burn have been observed. These tools hold significance in guiding healthcare policy and in communications with patients and their families.

## Data Availability

Contact the corresponding author for use of the data studied in this article.

## Declarations

### Ethical approval

IRB approval was obtained prior to the study. Disclaimer: Nothing to declare.

## REFERENCES

1. Abu-Hanna A, Lucas PJ. Prognostic models in medicine: AI and statistical approaches. Methods Inf Med. 2001;40:1–5

2. Hussain A, Dunn KW. Predicting length of stay in thermal burns: a systematic review of prognostic factors. Burns. 2013 Nov 1;39(7):1331–40.

3. Hussain A, Choukairi F, Dunn K. Predicting survival in thermal injury: a systematic review of methodology of composite prediction models. Burns. 2013 Aug 1;39(5):835–50.

4. Belgian Outcome Burn Injury Study Group. Development and validation of a model for prediction of mortality in patients with acute burn injury. Br J Surg 2009;96:111–7.

5. Tobiasen J, Hiebert JM, Edlich RF. The abbreviated burn severity index. Ann Emerg Med 1982;11:260–2.

6. Stern M, Waisbren BA. Comparison of methods of predicting burn mortality. Scand J Plast Reconstr Surg 1979;13:201–4.

7. Osler T, Glance LG, Hosmer DW. Simplified estimates of the probability of death after burn injuries: extending and updating the baux score. Journal of Trauma and Acute Care Surgery. 2010 Mar 1;68(3):690–7.

8. Gillespie R, Carroll W, Dimick AR, Haith L, Heimbach D, Kibbee E, et al. Diagnosis-related groupings (DRGs) and wound closure: roundtable discussion. J Burn Care Rehabil 1987; 8:199–209

9. Ho WS, Ying SY, Burd A. Outcome analysis of 286 severely burned patients: retrospective study. Hong Kong Med J 2002;8:235–9.

10. Akirov A, Masri-Iraqi H, Atamna A, Shimon I. Low albumin levels are associated with mortality risk in hospitalized patients. The American journal of medicine. 2017 Dec 1;130(12):1465–e11.

11. Feng JY, Chien JY, Kao KC, Tsai CL, Hung FM, Lin FM et al. Predictors of early onset multiple organ dysfunction in major burn patients with ventilator support: Experience from a mass casualty explosion. Scientific reports. 2018 Jul 19;8(1):1–9.

12. Frugoni B, Gabriel RA, Rafaat K, Abanobi M, Rantael B, Brzenski A. A predictive model for prolonged hospital length of stay in surgical burn patients. Burns. 2020 Nov 1;46(7):1565–70.

13. Klugman J. Human Development Report 2011. Sustainability and Equity: A better future for all. Sustainability and Equity: A Better Future for All (November 2, 2011). UNDP-HDRO Human Development Reports. 2011.

14. Zia N, Latif A, Mashreky SR, Al-Ibran E, Hashmi M, Rahman AF, Khondoker S, Quraishy MS, Hyder AA. Applying quality improvement methods to neglected conditions: development of the South Asia Burn Registry (SABR). BMC research notes. 2019 Dec;12(1):1–6.

15. Hussain J, Hanif A, Ali HA, Ashraf T. A Case Control Study with the Statistical Predictive Modeling of Child Burn in Pakistan. Journal of Statistics. 2013 Jan 1;20(1).

16. Zhang Z. Missing data imputation: focusing on single imputation. Annals of translational medicine. 2016 Jan;4(1).

17. Concato J, Feinstein AR, Holford TR. The risk of determining risk with multivariable models. Ann Intern Med 1993;118:201–10.

18. Meshulam-Derazon S, Nachumovsky S, Ad-El D, Sulkes J, Hauben DJ. Prediction of morbidity and mortality on admission to a burn unit. Plast Reconstr Surg 2006;118:116–20.

19. Sanderson LM, Buffler PA, Perry RR, Blackwell SJ. A multivariate evaluation of determinants of length of stay in a hospital burn unit. J Burn Care Rehabil 1981;2:142–9.

20. Saffle JR, Davis B, Williams P. Recent outcomes in the treatment of burn injury in the United States: a report from the American Burn Association Patient Registry. J Burn Care Rehabil 1995;16:219–32. discussion 88-9.

21. Smith DL, Cairns BA, Ramadan F, Dalston JS, Fakhry SM, Rutledge R, Meyer AA, Peterson HD. Effect of inhalation injury, burn size, and age on mortality: a study of 1447 consecutive burn patients. The Journal of trauma. 1994 Oct 1;37(4):655–9.

22. David GN, Micheal AG. Clinical features and diagnosis of lower extremity peripheral artery disease. UptoDate [Internet]. 2021 Feb 2. Available from: https://www.uptodate.com/contents/clinical-features-and-diagnosis-of-lower-extremity-peripheral-artery-disease

23. Pierpont YN, Dinh TP, Salas RE, Johnson EL, Wright TG, Robson MC, Payne WG. Obesity and surgical wound healing: a current review. International Scholarly Research Notices. 2014;2014.

24. Cantor AJ, Burger K. How to assess and manage burn injuries of the foot. Podiatry Today. 2004 July;17(7):46–52.

25. Wong MK, Ngim RC. Burns mortality and hospitalization time – a prospective statistical study of 352 patients in an Asian National Burn Centre. Burns 1995;21:39–46.

26. Bhatia AS, Mukherjee BN. Predicting survival in burned patients. Burns 1992;18:368–72

27. Mathers C. The global burden of disease: 2004 update. World Health Organization; 2008.

28. Michaud CM, Murray CJ, Bloom BR. Burden of disease—implications for future research. Jama. 2001 Feb 7;285(5):535–9.

